# Distinct respiratory tract biological pathways characterizing ARDS molecular phenotypes

**DOI:** 10.1101/2022.03.31.22272425

**Authors:** Aartik Sarma, Stephanie A. Christenson, Beth Shoshana Zha, Angela Oliveira Pisco, Lucile P.A. Neyton, Eran Mick, Pratik Sinha, Jennifer G. Wilson, Farzad Moazed, Aleksandra Leligdowicz, Manoj V. Maddali, Emily R. Siegel, Zoe M. Lyon, Hanjing Zhou, Alejandra Jauregui, Rajani Ghale, Saharai Caldera, Paula Hayakawa Serpa, Thomas Deiss, Christina Love, Ashley Byrne, Katrina L. Kalantar, Joseph L. DeRisi, David J. Erle, Matthew F. Krummel, Kirsten N. Kangelaris, Carolyn M. Hendrickson, Prescott G. Woodruff, COMET Consortium, Michael A. Matthay, Charles R. Langelier, Carolyn S. Calfee

## Abstract

**Background:** Two molecular phenotypes of the acute respiratory distress syndrome (ARDS) with divergent clinical trajectories and responses to therapy have been identified. Classification as “hyperinflammatory” or “hypoinflammatory” depends on plasma biomarker profiling. Differences in pulmonary biology underlying these phenotypes are unknown.

**Methods:** We analyzed tracheal aspirate (TA) RNA sequencing (RNASeq) data from 41 ARDS patients and 5 mechanically ventilated controls to assess differences in lung inflammation and repair between ARDS phenotypes. In a subset of subjects, we also analyzed plasma proteomic data. We performed single-cell RNA sequencing (scRNASeq) on TA samples from 9 ARDS patients. We conducted differential gene expression and gene set enrichment analyses, *in silico* prediction of pharmacologic treatments, and compared results to experimental models of acute lung injury.

**Findings:** In bulk RNASeq data, 1334 genes were differentially expressed between ARDS phenotypes (false detection rate < 0.1). Hyperinflammatory ARDS was characterized by an exaggerated innate immune response, increased activation of the integrated stress response, interferon signaling, apoptosis, and T-cell activation. Gene sets from experimental models of lipopolysaccharide lung injury overlapped more strongly with hyperinflammatory than hypoinflammatory ARDS, though overlap in gene expression between experimental and clinical samples was variable. ScRNASeq demonstrated a central role for T-cells in the hyperinflammatory phenotype. Plasma proteomics confirmed a role for innate immune activation, interferon signaling, and T-cell activation in the hyperinflammatory phenotype. Predicted candidate therapeutics for the hyperinflammatory phenotype included imatinib and dexamethasone.

**Interpretation:** Hyperinflammatory and hypoinflammatory ARDS phenotypes have distinct respiratory tract biology, which could inform targeted therapeutic development.

**Funding:** National Institutes of Health; University of California San Francisco ImmunoX CoLabs; Chan Zuckerberg Foundation; Genentech

## Introduction

The acute respiratory distress syndrome (ARDS) is characterized by noncardiogenic pulmonary edema and hypoxemia within one week of a physiologic insult^1^. The global incidence of ARDS has surged during the COVID-19 pandemic, increasing the importance of finding effective treatments. While some pharmacologic interventions have decreased mortality in patients with severe COVID-19^2,3^, no drug has consistently reduced mortality in more typical heterogeneous cohorts of patients with ARDS. There is a growing recognition that biological heterogeneity within the syndrome is a significant barrier to identifying effective treatments^4^.

Two clinically distinct molecular phenotypes of ARDS (termed “hyperinflammatory” and “hypoinflammatory”) have been identified using latent class analysis of clinical and plasma biomarker data in eight cohorts^1,5–11^. The hyperinflammatory phenotype is characterized by elevated plasma inflammatory cytokines (IL-8, IL-6, TNFr-1), lower plasma Protein C and bicarbonate, and higher mortality compared to the hypoinflammatory phenotype. Importantly, significant differences in treatment response to simvastatin, ventilator settings, and fluid management have been observed across molecular phenotypes in retrospective analyses of three ARDS clinical trials^5,6,11^; further, in patients with COVID-19-related ARDS, hyperinflammatory patients may preferentially respond to corticosteroid treatment^12,13^. These results suggest that understanding and targeting the heterogeneous biology underlying ARDS molecular phenotypes is essential to identifying effective new treatments for ARDS. Prospective studies designed to identify these phenotypes using parsimonious models are laying the groundwork for precision clinical trials^4,14^.

Despite this exciting progress, a critical barrier to developing new therapies for ARDS is the limited understanding of the biological pathways characterizing each phenotype. This knowledge gap was recently cited by NHLBI and European Respiratory Society workshops on precision medicine in ARDS as a top research priority for the field^4,15^. To date, analyses of the biological differences between these phenotypes have been largely limited to circulating biomarkers, due to the relative ease of sampling. Understanding the biological differences between ARDS molecular phenotypes in the lung, the active site of injury, will be critical to development of informative pre-clinical models of disease and targeted treatments for ARDS. Here, we employ a systems biology approach incorporating bulk and single-cell RNA-sequencing, *in silico* analyses, and proteomics to understand differences in lung immunology and inflammatory responses between ARDS phenotypes.

## Methods

### Study subjects

Subjects were enrolled in two prospective observational cohorts of critically ill patients. We used bulk RNASeq data and plasma proteomic data from the Acute Lung Injury in Critical Illness (ALI) study, a cohort of mechanically ventilated adults admitted to the intensive care unit at the University of California, San Francisco Medical Center (UCSFMC) between July 2013 and March 2020. We used single-cell RNA-Sequencing (scRNASeq) data from the COVID-19 Multiphenotyping for Effective Therapies (COMET) study, a study of hospitalized patients with COVID-19 or other acute respiratory illnesses admitted to UCSFMC or Zuckerberg San Francisco General Hospital (ZSFGH). COVID-19 status was confirmed by clinical PCR testing and metagenomic sequencing. These studies were approved by the UCSF Institutional Review Board (17-24056, 20-30497), which granted an initial waiver of informed consent to collect TA and blood samples. Informed consent was then obtained from patients or surrogates, as previously described^16^.

In this analysis, we included all available subjects in each cohort who were admitted to the intensive care unit for mechanical ventilation for ARDS or for airway protection without radiographic evidence of underlying pulmonary disease. For non-ARDS control patients in the ALI study, we excluded subjects on immunosuppression, including corticosteroids, and those with immunocompromising conditions (e.g., bone marrow transplant recipients).

### ARDS adjudication and phenotype assignment

Electronic health records were adjudicated for ARDS (Berlin Definition^17^) by at least two clinicians blinded to all biological data. Lower respiratory tract infections were diagnosed using the CDC surveillance definition^18^. ARDS phenotype was determined using a validated three-variable classifier model (IL-8, protein C, and bicarbonate)^14^. Subjects with a probability of class assignment greater than 0.5 were assigned to the hyperinflammatory phenotype. Plasma biomarkers were not available for five subjects with TA bulk RNA sequencing. For these subjects, we used a validated clinical classifier model to assign phenotype^10,19^.

### Tracheal aspirate sampling and RNA sequencing

Following enrollment in the ALI cohort, TA was collected within three days of intubation. In the COMET cohort, TA samples were collected daily, and the first available sample collected within three days of enrollment was used for this analysis. For bulk RNA sequencing, TA was collected and stored in DNA/RNA Shield (Zymo, Inc.) at -80C^16^. Samples underwent library preparation and Illumina paired-end sequencing using established methods described in detail in the online supplement. For scRNASeq, TA was collected and processed within 3 hours as previously described^20^.

### Cell annotation, differential expression, pathway, and network analysis

We performed pairwise comparisons of gene expression in each ARDS phenotype and controls using *DESeq2*. Single cell transcriptomes were annotated using *SingleR*. We compared gene expression between phenotypes for each cell type using *MAST*, using a mixed effects model with fixed effects for phenotype and cellular detection rate and a random effect for subject. Differentially expressed genes were then analyzed using Ingenuity Pathway Analysis (IPA, Qiagen). Full details of cell annotation, differential gene expression, and IPA analyses are provided in the online supplement. To study how cell-cell signaling contributed to observed differential expression, we used *CellChat*^21^ to infer intercellular communication networks by comparing scRNASeq data to a curated database of ligands, receptors, and their cofactors.

### Plasma proteomic analysis

Plasma samples from the Acute Lung Injury in Critical Illness cohort and 14 healthy controls from a previously published dataset^22^ were analyzed using the O-link Proteomics Assay, which generates a semi-quantitative measurement of 96 plasma proteins. We excluded all samples that were flagged with a QC warning from the O-link platform and excluded any biomarker for which protein concentrations could not be measured for at least 90% of samples. Measurements for 73 proteins passed the manufacturer’s quality control filter and were included for analysis. Normalized protein expression measurements were compared using a Wilcoxon rank-sum test and p-values were adjusted using the Benajmini-Hochberg method (FDR < 0.1).

### Comparison of differentially expressed genes to experimental models of acute lung injury

We identified experimental models of acute lung injury in the Gene Expression Omnibus. Lists of all genes differentially expressed at a Benjamini-Hochberg adjusted p-value less than

0.05 were downloaded using GEO2Enrichr^23^. We then used the genes that were upregulated in the experimental lung injury model as gene signatures in GSVA^24^ (Supplemental Data 3A). If more than 200 genes were differentially expressed in the experimental model, we used the top 200 genes (by p-value) for the experimental gene signature. We used *limma*^25^ to compare GSVA scores in samples from each phenotype to GSVA scores in controls.

## Results

### Patient Characteristics: Acute Lung Injury in Critical Illness Cohort

In the Acute Lung Injury in Critical Illness cohort, TA sequencing data was available for 41 ARDS participants and five controls (Figure S1). Ten of 41 ARDS subjects (24%) were classified as the hyperinflammatory phenotype, consistent with the proportion observed in previous studies^5–8,11^. There were no significant differences in age, sex, BMI, immunosuppression, respiratory viral or bacterial infections, or ARDS risk factors between hyperinflammatory and hypoinflammatory patients (Table 1).

**Table 1:** Characteristics of patients included in differential expression analysis of ARDS phenotypes from the Acute Lung Injury in Critical Illness cohort. Normally distributed values are reported as mean ± SD. Non-normally distributed values are reported as median [IQR]. Categorical data are reported as N (% of total for category). P-values are for a t-test for normally distributed continuous data, Wilcoxon rank-sum for non-normally distributed, and chi-square test for categorical data. P values are for (1) hyperinflammatory ARDS vs. hypoinflammatory ARDS; (2) hyperinflammatory ARDS vs. controls; (3) hypoinflammatory ARDS vs. controls

|  | Hyperinflammatory | Hypoinflammatory | P (1) | Control | P (2) | P (3) |
| --- | --- | --- | --- | --- | --- | --- |
| N | 10 | 31 |  | 5 |  |  |
| Age | 66 [56, 72] | 63 [51, 70] | 0.63 | 66 ± 23 | 0.85 | 0.42 |
| Female | 4 (40) | 21 (68) | 0.95 | 3 (60) | 0.26 | 0.49 |
| BMI (kg/m<sup>2</sup>) | 25.0 [23.6, 25.6] | 25.9 [23.9, 32.2] | 0.27 | 25.7 ± 4.8 | 0.24 | 0.23 |
| Vasopressors at enrollment | 9 (90) | 19 (61) | 0.19 | 1 (20) | <0.01 | 0.21 |
| Minimum PF ratio (mmHg) | 76 [62, 97] | 93 [67, 138] | 0.25 | 296 [216,366] | <0.01 | <0.01 |
| SOFA score at enrollment | 18 [16, 19] | 9 [7, 11] | <0.01 | 5 [4, 5] | <0.01 | <0.01 |
| IL-8, pg/ml | 424 [228, 1068] | 15 [9, 25] | <0.01 | 10 [8, 11] | <0.01 | 0.21 |
| Protein C, % control | 51 [31, 62] | 103 [76, 132] | <0.01 | 115 [79, 148] | <0.01 | 0.76 |
| Immunosuppression | 4 (40) | 6 (19) | 0.37 |  |  |  |
| Primary ALI Risk Factor |  |  | 0.79 |  |  |  |
| *Pneumonia* | 4 (40) | 16 (48) |  |  |  |  |
| *Sepsis* | 4 (40) | 7 (23) |  |  |  |  |
| *Aspiration* | 2 (20) | 6 (23) |  |  |  |  |
| *Pancreatitis* | 0 (0) | 1 (3) |  |  |  |  |
| *None* | 0 (0) | 1 (3) |  |  |  |  |
| **Clinical respiratory microbiology** |  |  |  |  |  |  |
| *Respiratory viral pathogen* | 1 (10) | 1 (3) | 0.98 |  |  |  |
| *Respiratory bacterial pathogen* | 3 (30) | 11 (36) | 1.00 |  |  |  |

### Bulk RNA-Sequencing

1,334 genes were differentially expressed between ARDS phenotypes with an absolute empirical Bayesian posterior log_2_-fold change >0.5 (Figure S2A, Supplementary Data S1A). IPA predicted increased activation of several cytokines and other upstream regulators of differentially expressed genes in hyperinflammatory ARDS, compared with hypoinflammatory ARDS (Figure 1A, Supplementary Data S2A). These included several cytokines classically associated with an innate response previously found to be elevated in plasma of patients with hyperinflammatory ARDS, including IL1B, IL6, and TNF. In addition, IPA identified activation of cytokine responses not previously studied in hyperinflammatory ARDS, including numerous interferon-stimulated genes; IL2 and IL15, which stimulate cytotoxic T cell and NK cell responses^26^; and the chemokine ligand CCL2/MCP-1. Upstream regulator analysis predicted increased activation of transcriptional regulators critical to the integrated stress response (XBP1, NFE2L2) and increased cellular differentiation (MYC, NONO), as well as stimulation of Toll-like receptors (TLR2, TLR3, TLR4, TLR7, TLR9) and receptors integral to T cell activation (CD3, CD28) in hyperinflammatory ARDS.

**Figure 1:**
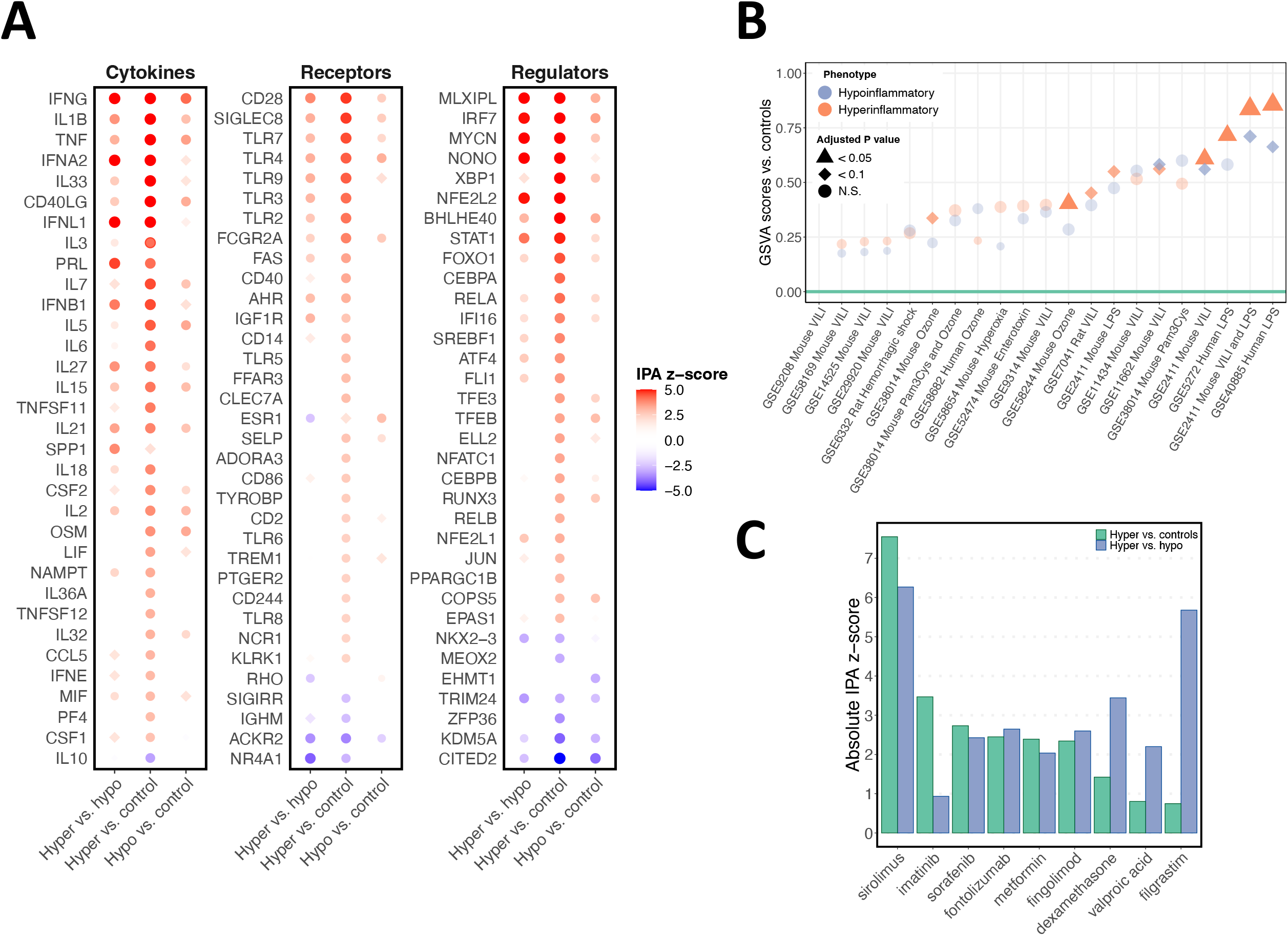
Bulk RNA sequencing analyses of TA collected in the Acute Lung Injury in Critical Illness cohort. **(A)** Upstream regulator z-scores based on IPA analysis of differential gene expression in pairwise comparisons of tracheal aspirate bulk RNA sequencing in hyperinflammatory ARDS (N=10), hypoinflammatory ARDS (N=31), and mechanically ventilated controls (N=5). A positive z-score indicates differential gene expression is consistent with greater activation of the upstream regulator of gene expression in the first group in each pairwise comparison. Circles identify upstream regulators that are statistically significant. **(B)** Gene set variation analysis for experimental models of lung injury. GSVA scores were calculated for each sample, and the difference between hyperinflammatory ARDS and controls (orange) and hypoinflammatory ARDS and controls (blue) was estimated with limma. For each model, the GEO Accession Number, organism, and lung injury model are listed on the x-axis. (**C)** Upstream regulator scores for selected drugs in the IPA database that are predicted to significantly shift gene expression away from hyperinflammatory ARDS in independent analyses of differential gene expression with hypoinflammatory ARDS (blue) or controls (green).

To further understand how pathways in each phenotype were dysregulated, we compared each phenotype to mechanically ventilated control patients with neurologic injury. 2,989 genes were differentially expressed between hyperinflammatory ARDS and controls (Figure S2B, Supplementary Data 1C), while 2,132 genes were differentially expressed between hypoinflammatory ARDS and controls (Figure S2C, Supplementary Data 1D). Notably, upstream regulator analysis identified several cytokines that were activated in both hyperinflammatory and hypoinflammatory ARDS compared to controls (Figure 1C, Supplementary Data 2C and 2D), including IL1B, TNF, and IFNG. While this analysis identified some similarities between phenotypes, it also confirmed that several upstream regulators that were activated in pairwise comparisons of hyperinflammatory ARDS to hypoinflammatory ARDS and hyperinflammatory ARDS to controls, suggesting these upstream regulators play an important role in the distinct features of hyperinflammatory ARDS. These included IL6, which was one of plasma cytokines used to define the hyperinflammatory phenotype^5^; IL18, which was not measured in the original LCA studies, but, like the hyperinflammatory phenotype, was associated with higher mortality and a response to simvastatin in the HARP-2 trial^27^; the T cell receptor; Type I/III interferons (IFNA, IFNL, IFNB), indicative of an enhanced mucosal interferon response^28^; several Toll-like receptors (TLR2/3/9); and FAS, which stimulates apoptosis^29^. In addition, several upstream regulators associated with specific immune cells were predicted to be activated in hyperinflammatory ARDS but not in hypoinflammatory ARDS, including markers of activated T cells (NFATC1^30^), NK cells (NCR1^31^, KLRK1^32^), and platelets (PF4^33^), suggesting these cells play a key role in the distinct biology of hyperinflammatory ARDS.

### Alignment with Experimental Models of Lung Injury

IPA identified lipopolysaccharide (LPS), a component of gram-negative bacteria, as an upstream regulator of genes differentially expressed between ARDS phenotypes (Supplementary Data S2A) and in comparisons of each ARDS phenotype to controls (Supplementary Data S2C and S2D). We hypothesized that genes upregulated in LPS models of lung injury would be more upregulated in hyperinflammatory ARDS compared to controls than in hypoinflammatory ARDS compared to controls. Respiratory tract gene expression data was available from four LPS models of ARDS in the Gene Expression Omnibus. We also identified 17 more datasets from other experimental models of ARDS including ventilator-induced lung injury (VILI), ozone, hyperoxia, Pam3Cys (a TLR2 agonist), and hemorrhagic shock (Supplementary Data S3A). Gene sets from four models were significantly enriched (FDR < 0.1) in TA from both ARDS phenotypes (Figure 1B; Supplementary Data S3B and S3C). As expected, LPS models had a significant overlap with both phenotypes, but LPS experimental gene sets had higher GSVA scores in hyperinflammatory participants. In addition, gene sets from two ozone models, two LPS models, and one VILI model were enriched in hyperinflammatory ARDS but not in hypoinflammatory ARDS, suggesting these models better replicated dysregulated gene expression observed in the hyperinflammatory phenotype.

### *In silico* analysis of candidate drugs for hyperinflammatory ARDS

To identify candidate treatments for the hyperinflammatory ARDS phenotype, we used Upstream Regulator Analysis to identify drugs predicted to decrease expression of genes upregulated in hyperinflammatory ARDS compared to hypoinflammatory ARDS or controls (Figure 1C). For example, dexamethasone, which decreases interferon-gamma signaling^34^, was predicted to shift gene expression away from hyperinflammatory ARDS. Interestingly, several drugs which cause drug-induced pneumonitis (e.g., nitrofurantoin, amiodarone, and cytarabine) were predicted to shift gene expression from controls toward hyperinflammatory ARDS (Supplementary Data S2C).

### Single-cell RNA-sequencing

We used a neutrophil-preserving scRNASeq method to study TA from nine COVID-negative patients with ARDS enrolled in a separate observational cohort (COMET; described in Methods). TA scRNASeq was available from five participants with hypoinflammatory ARDS and four with hyperinflammatory ARDS; clinical characteristics of these patients are provided in Supplementary Table 1. 26,429 cells passed quality filters, and we used SingleR^35^ to identify cell types (Figure 2A). Neutrophils were the most common cell in both phenotypes (Figure 2B),

**Figure 2:**
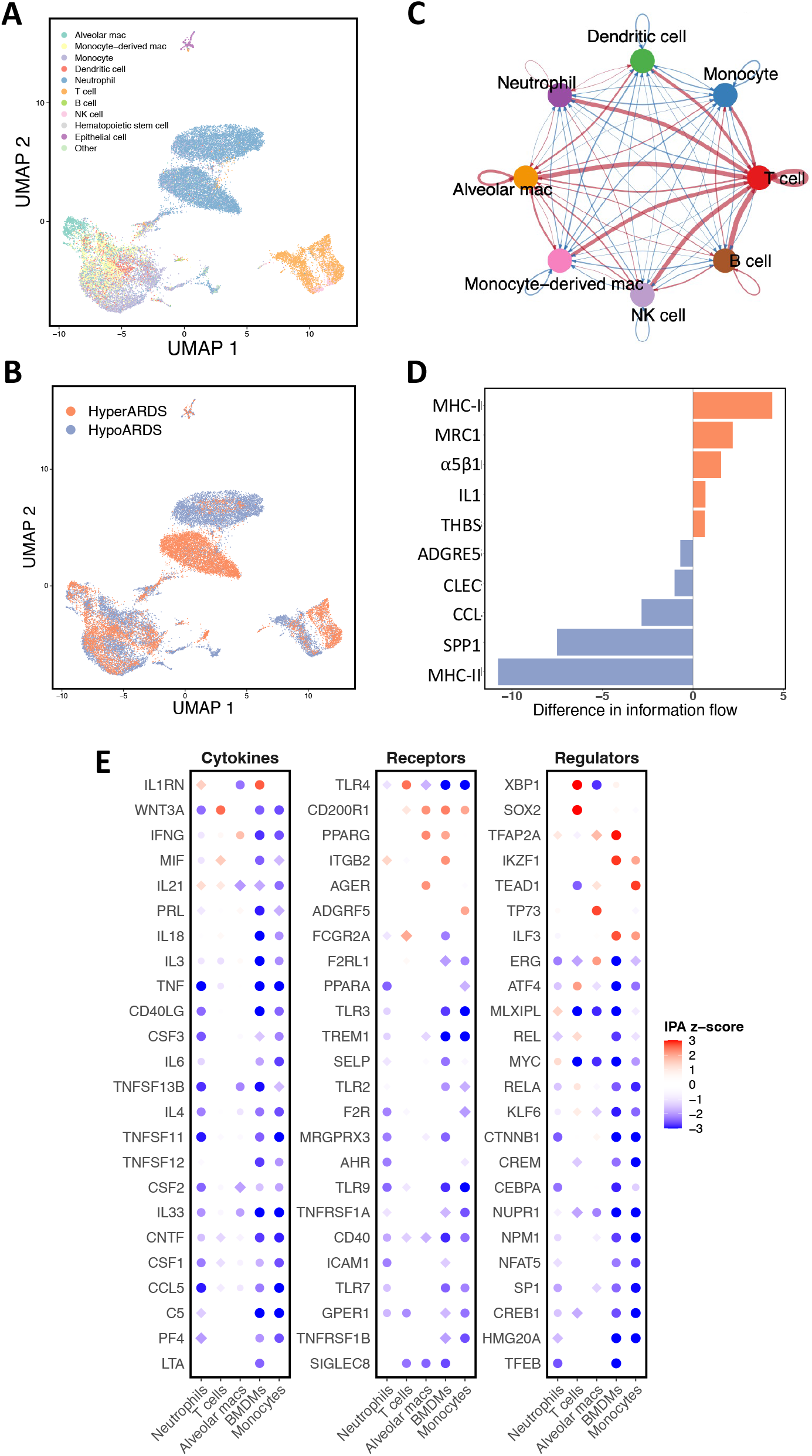
TA single-cell RNA sequencing. **(A)** Seurat UMAP projection of 26,429 TA cell transcriptomes from four participants with hyperinflammatory ARDS and five participants with hypoinflammatory ARDS, annotated with cell type as predicted by SingleR. **(B)** UMAP projection of TA cells transcriptomes separated by ARDS phenotype. **(C)** Differential interaction between cell types predicted by CellChat. Red arrows identify cell pairs with greater strength of interaction in hyperinflammatory ARDS. **(D)** Differences in strength of ligand-receptor interaction for pathways in the CellChat database. **(E)** Upstream regulator analysis z-scores for differentially expressed genes in neutrophils, monocytes, monocyte-derived macrophages, and T cells. A positive z-score indicates the upstream regulator is predicted to be more highly activated in hyperinflammatory ARDS. Circles identify upstream regulators that were statistically significant.

*CellChat* predicted markedly higher interaction between T cells and other cell types in the hyperinflammatory phenotype (Figure 2C), which was consistent with the bulk analysis predicting increased T cell activation. Phenotype-specific differences in cell-cell signaling were driven by ligand-receptor pairs in several pathways (Figure 2D; Supplementary Figure 6A and 6B). In the hyperinflammatory phenotype, *CellChat* predicted increased MHC-I signaling, which was driven by increased signaling to CD8 on T cells and NK cells. In contrast, MHC-II activity was predicted to be higher in hypoinflammatory ARDS. *CellChat* also identified increased NAMPT signaling by NK and T cells to integrin L5β1 on monocytes, macrophages, and dendritic cells in hyperinflammatory ARDS (Supplementary Data 6). Notably, NAMPT polymorphisms are associated with a 7.7-fold increased risk of sepsis-associated ARDS^36^ and NAMPT was identified as an upstream regulator of differential gene expression in our bulk RNASeq data (Figure 1C). These results further supported the hypothesis that there are marked differences in respiratory tract signaling between phenotypes driven by differences in T and NK cell signaling.

We next used *MAST* to compare differential gene expression in TA neutrophils, T cells, monocyte-derived macrophages, and monocytes (Supplementary Figure 4, Supplementary Data S4). We used IPA to identify upstream regulators of gene expression and analysis identified several cell-specific differences in gene expression (Figure 2E, Supplementary Data S5). Notably, some cytokines that were predicted to be activated in hyperinflammatory ARDS in the bulk RNA sequencing data, including TNF and IFNG, were relatively less active in neutrophils and monocytes from hyperinflammatory TA samples. In contrast, interferon lambda, which is produced by the respiratory epithelium, was predicted to be more activated in hyperinflammatory neutrophils and T cells. IPA also predicted increased activation of cell activation of the integrated stress response (XBP1, EIF2AK2) and TLR4 in T cells from the hyperinflammatory phenotype (Supplementary Data S5D).

### Plasma proteomic analysis identifies additional cytokines upregulated in hyperinflammatory ARDS

To further validate the biologic relevance of the TA findings, we measured plasma concentrations of 96 protein biomarkers. 21 participants included in the TA bulk sequencing analysis had protein biomarker data available, as did four participants from the same cohort who did not have TA bulk sequencing available. Of these 25 participants, five had hyperinflammatory ARDS and 20 had hypoinflammatory ARDS (Supplementary Table 2). We compared these subjects to 14 healthy controls (mean age: 38, 43% female) and included in a previously published analysis^22^.

Plasma concentrations of 28 proteins were higher in hyperinflammatory ARDS than in hypoinflammatory ARDS (FDR < 0.1, Figure 3A). Some of these biomarkers confirmed known differences between phenotypes, including higher concentrations of IL6 and TNF in hyperinflammatory ARDS. Nine of these biomarkers were also elevated in hyperinflammatory ARDS compared to controls but were not elevated in hypoinflammatory ARDS (Figure 3B and 3C), suggesting they identify distinctly dysregulated pathways in the hyperinflammatory phenotype. These proteins included IL-8, which is one of the cytokines that defines the hyperinflammatory phenotype; CASP-8, an effector of FAS signaling^37^; interferon-induced proteins CXCL9 and CXCL10^38^; plasma urokinase (uPA); oncostatin M; and adenosine deaminase (ADA). In addition, CCL2/MCP-1 and the T cell activation marker CD5^39^ were higher in hyperinflammatory ARDS and in controls compared to hypoinflammatory ARDS. These observations were consistent with differences in TA gene expression at both the bulk RNASeq and scRNASeq level. Plasma proteins that were higher in controls than in ARDS subjects are shown in Supplementary Figure S7.

**Figure 3:**
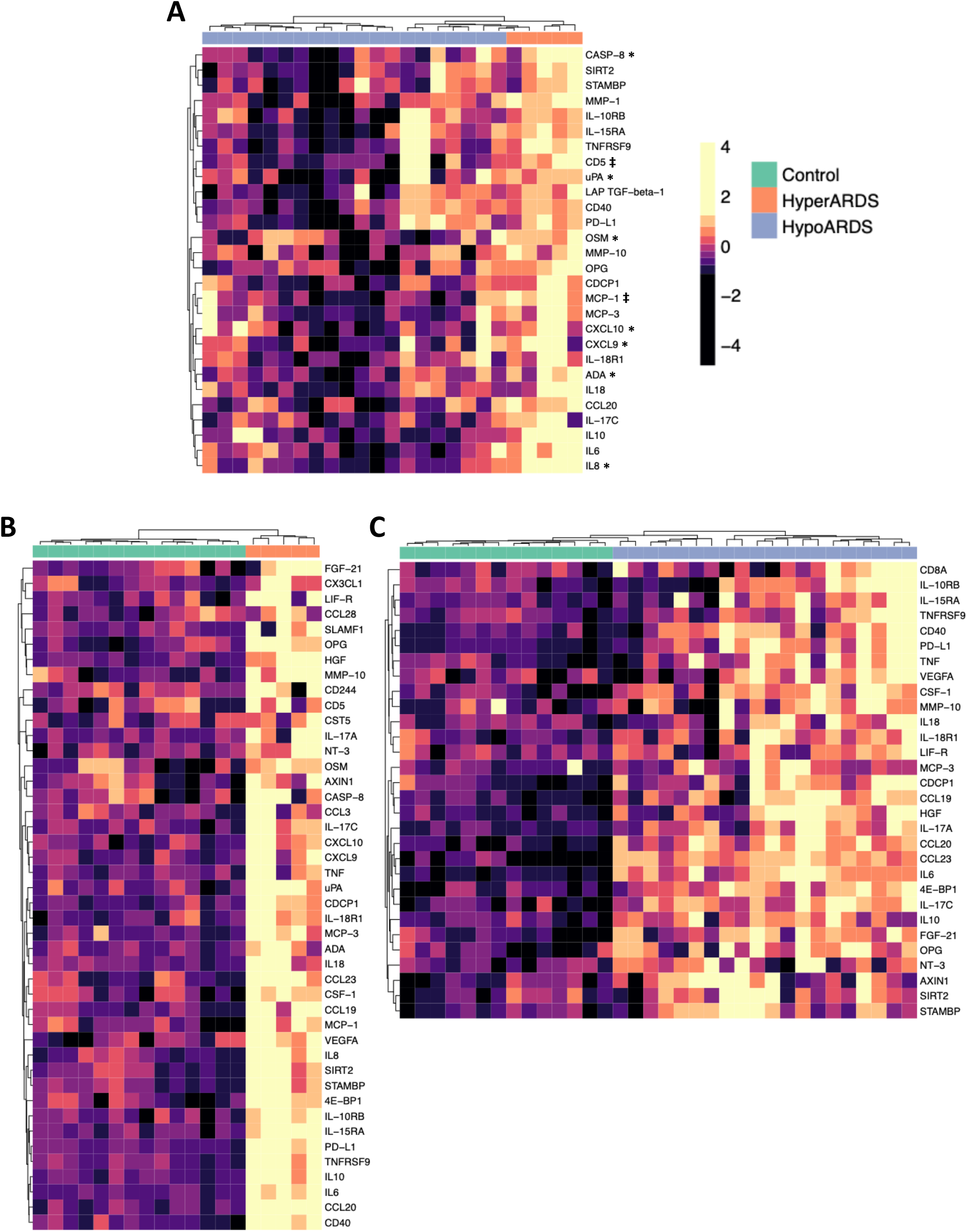
O-Link proteomics results for plasma biomarkers from 5 hyperinflammatory ARDS, 20 hypoinflammatory ARDS, and 14 control participants. Each heatmap shows plasma protein biomarkers that were significantly different between groups (FDR < 0.1); for a complete list of proteins, see Supplementary Data S7. Each column represents an individual subject, and each row shows the z-scaled concentrations. Rows and columns are clustered using the Euclidean distance. Columns are annotated by phenotype. Z-score for expression is shown on the color bar on the right. **(A)** Hyperinflammatory ARDS vs. hypoinflammatory ARDS (*: proteins that higher in hyperinflammatory ARDS vs. volunteers but not difference between hypoinflammatory ARDS and controls; ‡: proteins that are higher in hyperinflammatory ARDS vs. volunteers and lower in hypoinflammatory ARDS vs. volunteers) **(B)** Hyperinflammatory ARDS vs. healthy volunteers. **(C)** Hypoinflammatory ARDS vs. healthy volunteers.

## Discussion

This analysis represents the first report of significant differences in pulmonary biology between ARDS molecular phenotypes, which have previously been characterized primarily using plasma biomarkers. In addition to confirming evidence of innate immune activation suggested by prior data on circulating plasma biomarkers, these analyses identify several novel pathways as relevant to the pathogenesis of hyperinflammatory ARDS, including interferon-stimulated pathways, apoptosis, and T-cell signaling, and suggest that each ARDS phenotype has distinct pulmonary pathobiology which could help identify new therapeutic targets.

We identified a central role of T and NK cells in coordinating dysregulated inflammation in hyperinflammatory ARDS. Hyperinflammatory ARDS was associated with markedly higher mucosal interferon-stimulated gene expression and T/NK cell activation in bulk sequencing of TA. Network analysis of scRNASeq data was also consistent with a central role of T and NK cells in hyperinflammatory ARDS. These analyses identified differences in APC to T/NK cell signaling, with greater MHC-I to CD8 signaling in the hyperinflammatory phenotype. Single cell differential expression also predicted increased activation of TLR4 and XBP1 in hyperinflammatory ARDS, suggesting the integrated stress and innate immune responses are upregulated in T cells from the hyperinflammatory phenotype. In contrast to prior literature on ARDS^40,41^, which has emphasized the role of neutrophils, activated macrophages, and alveolar epithelial cells in ARDS pathogenesis, our observations suggest lymphocytes play an underrecognized role in coordinating dysregulated inflammation in hyperinflammatory ARDS.

Some features that characterized the hyperinflammatory phenotype in bulk sequencing (e.g., higher predicted activation of cytokines classically associated with an innate response) were not observed in specific cell types in scRNASeq. This observation has several possible explanations. First, the results in bulk RNA sequencing may be driven by differences in the immune cell composition of TA in each phenotype, as suggested by our scRNASeq analyses. Notably, a similar pattern of high interferon-stimulated gene expression in T cells but diminished immune responses in macrophages has also been reported in severe COVID-19^42^. Second, the bulk signals may be driven by highly activated cells that do not survive the scRNASeq processing pipeline, as has been reported previously for activated macrophages and neutrophils. Third, some signals observed in the bulk data may be driven by epithelial cells, which were selected against in our scRNASeq pipeline (in favor of enriching for immune cell populations) and are thus not well-represented.

Plasma proteomics identified increased concentrations of the interferon-stimulated proteins CXCL9 and CXCL10 and the T cell activation marker CD5 in hyperinflammatory ARDS but not in hypoinflammatory ARDS. Notably, in an alternative molecular phenotyping approach that used *k-*means clustering of plasma biomarkers to categorize ARDS subjects into two molecular phenotypes (“reactive” and “uninflamed”), plasma IFNγ is one of the defining biomarkers of the higher mortality “reactive” phenotype^43^. Taken together, these analyses support a central role of mucosal interferons and T cell activation in hyperinflammatory ARDS.

To our knowledge, only one prior study has attempted to characterize the pulmonary compartment in ARDS phenotypes^44^. This study included 10 hypoinflammatory patients and 16 hyperinflammatory patients and found no difference between these groups in concentration of several inflammatory protein biomarkers in mini-BAL samples or in the lung microbiome. Our results may differ from these because of a larger sample size, differing analytic approaches, differing sampling strategies, or some combination thereof.

We compared differentially expressed genes in clinical samples to experimental ARDS models to determine the relevance of these models to each phenotype. An experimental model combining intratracheal LPS *and* mechanical ventilation was the murine model with the strongest overlap in gene expression with ARDS subjects in both phenotypes. This gene signature was from an experiment demonstrating that a combined MV/LPS model generated markedly higher neutrophilic inflammation in the lung than LPS or MV alone^45^. In addition, gene signatures from five LPS models were enriched in hyperinflammatory ARDS but were not enriched in hypoinflammatory ARDS, suggesting that hypoinflammatory patients, on average, have less of an overlap in respiratory biology with the preclinical models. Our results also indicate that the overlap in gene expression between experimental models and clinical samples is highly variable, even among models with similar injurious stimuli (e.g., among VILI models). Further study is required to understand how best to model each phenotype experimentally and if our observations partially explain why therapies that appear promising in pre-clinical models are not effective in more heterogeneous clinical trial populations.

Our results have important implications for developing a precision approach to treating ARDS^4^. The *in silico* analysis identified several candidate therapies that target the dysregulated pathways identified in hyperinflammatory ARDS. Several of the candidate drugs, including imatinib, dexamethasone, and metformin, decrease lung injury caused by LPS in experimental models^46–48^, again suggesting LPS replicates important features of the hyperinflammatory phenotype. Approximately one-third of candidate drugs identified using pathway analyses are validated in *in* vivo^49^ and these treatments require validation in further preclinical studies and clinical trials.

Strengths of this study include transcriptomic analysis of samples from the focal organ of injury in ARDS, providing a detailed picture of the pulmonary biology of both ARDS phenotypes, and deepening of these observations with single-cell sequencing and peripheral blood proteomics. The inclusion of non-ARDS ventilated controls allowed us to further characterize the physiologic dysregulation in the phenotypes, rather than defining gene expression relative to another pathologic state. This analysis also has some limitations. Bulk and single-cell RNASeq samples were collected in separate cohorts, so we cannot integrate these analytical approaches. Although TA contains fluid from the distal airspaces^50^, more invasive BAL testing may identify additional differences between the phenotypes.

In conclusion, an integrated, multi-omic analysis of ARDS molecular phenotypes originally defined by clinical and plasma protein biomarkers suggests the hyperinflammatory phenotype is characterized by increased interferon-stimulated gene expression coordinated by T cell signaling in the lower respiratory tract. Our findings suggest that the respiratory tract biology of these phenotypes is distinct and further supports the use of molecular phenotypes to study acute lung injury biology and develop new treatments for ARDS.

## Materials and Methods

## Data Availability

All data produced in the present study are available upon reasonable request to the authors

## List of Supplementary Materials

**Figure S1:** Subjects included in this analysis.

**Figure S2:** Volcano plots showing differential gene expression between (A) 10 hyperinflammatory ARDS and 31 hypoinflammatory ARDS subjects, (B) 10 hyperinflammatory ARDS and 5 control subjects, and (C) 31 hypoinflammatory ARDS and 5 control subjects.

**Figure S3:** GSVA scores for signatures of ALI experimental models in TA transcriptomes from 5 control subjects, 31 hypoinflammatory ARDS subjects, and 10 hyperinflammatory ARDS subjects.

**Figure S4:** Volcano plots showing differential gene expression in scRNASeq for 5 hypoinflammatory ARDS subjects and 4 hypoinflammatory ARDS subjects in the COMET cohort.

**Figure S5**: Circos plots displaying differentially interactive ligand-receptor pairs identified by CellChat in a comparison of scRNASeq from 5 hypoinflammatory and 4 hyperinflammatory ARDS subjects. (A) Pairs that are relatively higher in hyperinflammatory ARDS and (B) Pairs that are relatively higher in hypoinflammatory ARDS.

**Figure S6**: Heatmaps for plasma proteins that are significantly higher in (A) Controls vs. hyperinflammatory ARDS and (B) Controls vs. hypoinflammatory ARDS

**Figure S7**: Heatmap showing estimated power to detect a difference between phenotypes in the Acute Lung Injury in Critical Illness cohort based on effect size and within-group coefficient of variation.

**Supplementary Data S1:** Differential gene expression for pairwise comparisons of bulk RNA gene expression for TA in the Acute Lung Injury in Critical Illness cohort. A positive log_2_ fold difference indicates the gene is more highly expressed in the first group compared to second group for each comparison. (A) Hyperinflammatory ARDS vs. hypoinflammatory ARDS; (B) Hyperinflammatory ARDS vs. hypoinflammatory ARDS after adjusting for fungal infections; (C) Hyperinflammatory ARDS vs. controls; and (D) Hypoinflammatory ARDS vs. controls; and (E) All ARDS subjects vs. controls.

**Supplementary Data S2:** IPA Upstream Regulator scores for pairwise comparisons of bulk RNA gene expression for TA in the Acute Lung Injury in Critical Illness Cohort. A positive z-score indicates gene expression is consistent with higher activity of the upstream regulator in the first group compared to second group for each comparison. (A) Hyperinflammatory ARDS vs. hypoinflammatory ARDS; (B) Hyperinflammatory ARDS vs. controls; (C) Hypoinflammatory ARDS vs. controls; and (D) All ARDS vs. controls.

**Supplementary Data S3**: (A) 200 most upregulated genes in experimental models of lung injury compared to controls for 21 experimental systems in the Gene Expression Omnibus. (B) Gene set enrichment analysis scores and leading-edge genes for experimental model gene sets in TA differential expression for hyperinflammatory ARDS vs. controls. (C) Gene set enrichment analysis scores and leading-edge genes for experimental model gene sets in TA differential expression for hypoinflammatory ARDS vs. controls.

**Supplementary Data S4:** Differential gene expression for single-cell RNA sequencing for TA for the COMET cohort. A positive log_2_ fold difference indicates the gene is more highly expressed in hyperinflammatory ARDS. (A) Neutrophils, (B) Monocytes, (C) Monocyte-derived macrophages, (D) T cells

**Supplementary Data S5:** IPA Upstream Regulator scores for pairwise comparisons of bulk RNA gene expression for TA in the COMET. A positive z-score indicates gene expression is consistent with higher activity of the upstream regulator in the hyperinflammatory ARDS samples. (A) Neutrophils, (B) Monocytes, (C) Monocyte-derived macrophages, (D) T cells

**Supplementary Data S6:** CellChat results for ligand-receptor pairs in TA scRNASeq from the COMET cohort for (A) hyperinflammatory ARDS and (B) hypoinflammatory ARDS.

**Supplementary Data S7: (**A) Z-scaled O-link protein concentrations for five hyperinflammatory ARDS, 20 hypoinflammatory ARDS, and 14 control subjects. (B) Results of Wilcoxon rank-sum tests for pairwise comparisons of hyperinflammatory ARDS, hypoinflammatory ARDS, and control subjects.

## Funding

National Institutes of Health grant F32HL151117 (AS)

National Institutes of Health grant R35HL140026 (CSC)

National Institutes of Health grant 2R24AA019661-06A1 (CSC)

National Institutes of Health grant K23HL138461-01A1 (CRL)

National Institutes of Health grant U19AI1077439 (DJE, CSC)

University of California San Francisco ImmunoX CoLabs

Chan Zuckerberg Foundation 2019-202665

Genentech TSK-020586

## Author contributions

Conceptualization: AS, SAC, CSC

Formal analysis/software: AS, SAC, AOP, LPAN, EM, MVM, AB, KLK

Data curation: HZ, AJ, RG, SC, TD

Investigation: AS, BSZ, PS, JGW, FM, AL, ERS, ZML, RG, SC, PHS, TD, CL, KNK, CMH, MAM, CRL, CSC

Visualization: AS

Funding acquisition: AS, CRL, DJE, CSC

Project administration: JLD, DJE, MFK, KNK, CMH, PGW, MAM, CRL, CSC

Supervision: SAC, CRL, CSC

Writing – original draft: AS

Writing – review & editing: All authors

## Data and materials availability

Code for differential expression and scRNASeq analyses is available at https://github.com/AartikSarma/ARDSPhenotypes

The processed gene count data are available from the National Center for Biotechnology Information Gene Expression Omnibus database under accession code GSE200849. The raw sequencing data used in this analysis are protected due to data privacy restrictions from the IRB protocols governing patient enrollment in this study, which protect the release of raw genetic sequencing data from those patients enrolled under a waiver of consent. Researchers who wish to obtain raw FASTQ files for the purposes of independently generating gene counts can contact the corresponding author to be added to the IRB protocols and sign a materials transfer agreement from UCSF ensuring that the data will be securely stored and only utilized for transcriptomic analyses.

