## Supplementary Methods for "Distinct respiratory tract biological pathways characterizing ARDS molecular phenotypes"

**Bulk RNA sequencing**: Following RNA extraction (Zymo Pathogen Magbead Kit) and DNase treatment, human cytosolic and mitochondrial ribosomal RNA was depleted using FastSelect (Qiagen). To control for background contamination, we included negative controls (water and HeLa cell RNA) as well as positive controls (spike-in RNA standards from the External RNA Controls Consortium (ERCC)). RNA was fragmented and underwent library preparation using the NEBNext Ultra II RNASeq Kit (New England Biolabs). Libraries underwent 146 nucleotide paired-end sequencing on an Illumina Novaseq 6000 instrument.

Following demultiplexing, sequencing reads were pseudo-aligned with *kallisto* (v. 0.46.1) to an index consisting of all transcripts associated with human protein coding genes (ENSEMBL v. 99), cytosolic and mitochondrial ribosomal RNA sequences. Gene-level counts were generated from the transcript-level abundance estimates using the R package *tximport*, with the scaledTPM method. Samples retained in the dataset had a total of at least 500,000 estimated counts associated with transcripts of protein coding genes, and the median across all samples was 7,528,890 counts. We reviewed PCA plots, hierarchical clustering, and percentage of non-zero reads per sample for quality control and excluded four bulk RNASeq samples as technical outliers.

**Analysis of differential gene expression, bulk RNA sequencing data:** Gene expression was fit to a model using the design formula ~Seqeuncing Batch + Gender + Z-scaled Age + Diagnosis. We performed pairwise comparisons of 1) hyperinflammatory ARDS to hypoinflammatory ARDS, 2) all ARDS subjects to mechanically ventilated controls, 3) hyperinflammatory ARDS to controls, and 4) hypoinflammatory ARDS to controls. Empirical Bayesian shrinkage estimators for log_2_-fold change were fit using *apeglm*. Significant differentially expressed genes were identified by an independent-hypothesis weighted false detection rate (FDR) < 0.1 and absolute shrunken log_2_FoldChange greater than 0.5. We estimated the power to detect a difference between groups at an alpha of 0.1 and average read depth of 100 across a biologically plausible range of within-group coefficients of variation using the RNASeqPower library for R, which confirmed our sample was adequately powered to detect large changes in gene expression between phenotypes (Supplementary Figure S8).

**Ingenuity Pathway Analysis (IPA), additional methods:** IPA (Qiagen, Inc.) compares differentially expressed genes to a database of gene signatures derived from experimental datasets. In addition to identifying signatures of canonical pathways and cellular function, IPA includes a database of upstream regulators of gene function. These include endogenous signaling molecules, such as cytokines, and exogenous stimuli, such as drugs or toxins. The dataset also includes annotations of the relationship between regulators, which can be used to construct mechanistic networks of regulators that may indirectly affect gene expression. For each of the signatures, IPA calculates a p-value for the overlap of DE genes with genes in the signature (using Fisher’s exact test) and a z-score that tests whether the measured direction of gene expression is consistent the direction in a gene signature. We defined significant pathways, regulators, and networks as those with a Benjamini-Hochberg false detection rate less than 0.1 or an absolute z-score greater than 2.

**Single cell RNA sequencing, additional methods**

TA cells were isolated and selected for expression of CD45, prepared with a V(D)J v1.1 kit according to the manufacturer’s protocol, processed on a 10x Genomics Chip A without multiplexing, and sequenced on an Illumina NovaSeq 6000 platform. Because samples had to be processed shortly after collection, batch processing and multiplexing were not feasible. Transcripts were aligned in Cell Ranger and filtered count matrices were imported into a Seurat object. Cells were filtered to have at least 300 counts, no more than 30,000 counts, and less than 10% mitochondrial genes.

Single-cell transcriptomes were assigned cell labels by SingleR, which compares single cell gene expression to a reference dataset (Human Primary Cell Atlas). Cell-cell communication networks in each phenotype were inferred using *CellChat,* which compares single cell gene expression to a curated database of ligand-receptor pairs. Differential gene expression between ARDS phenotypes was estimated using mixed effects models in *MAST*, with a random effect for subject and fixed effect for phenotype, adjusted for number of counts in each cell.

**Plasma proteomic analysis, additional methods**

Plasma samples were analyzed using the O-link Proteomics Assay, which generates a semi-quantitative measurement of 96 plasma proteins. We excluded all samples that were flagged with a QC warning from the O-link platform and excluded any biomarker for which protein concentrations could not be measured for at least 90% of samples. Measurements for 73 proteins passed the manufacturer’s quality control filter and were included for analysis.
