## Supplementary Data 8 for "Distinct respiratory tract biological pathways characterizing ARDS molecular phenotypes"

|  | **Hyperinflammatory** | **Hypoinflammatory** |  | P |
| --- | --- | --- | --- | --- |
| **N** | 4 | 5 |  |  |
| **Age** | 62 [57, 62] | 65 [49, 77] |  | 0.33 |
| **Female** | 2 (50) | 3 (60) |  | 1.00 |
| **BMI (kg/m^2^)** | 24.1 [21.1, 28,5] | 295 [27.8, 39.0] |  | 0.09 |
| **Vasopressors at enrollment** | 4 (100) | 4 (80) |  | 1.00 |
| **Minimum PF ratio (mmHg)** | 98 [78, 130] | 200 [138, 236] |  | 0.14 |
| **SOFA score at enrollment** | 19 [17, 20] | 9 [8, 12] |  | 0.01 |
| **IL-8, pg/ml** | 164 [94, 393] | 33 [3, 43] |  | 0.09 |
| **Protein C, % control** | 16 [15, 19] | 70 [61, 169] |  | 0.03 |
| **Immunosuppression** | 0 (0) | 1 (20) |  | 1.00 |
| **Primary ALI Risk Factor** |  |  |  | 0.79 |
| *Pneumonia* | 3 (75) | 3 (60) |  |  |
| *Sepsis* | 1 (25) | 0 (0) |  |  |
| *Aspiration* | 0 (0) | 1 (20) |  |  |
| *Pancreatitis* | 0 (0) | 1 (20) |  |  |
| **Clinical microbiology** |  |  |  |  |
| *Respiratory bacterial pathogen* | 1 (25) | 2 (40) |  | 1.00 |
| *Extrapulmonary virus* | 1 (25) | 0 (0) |  | 0.91 |
| *Extrapulmonary bacteria* | 3 (75) | 0 (0) |  | 0.10 |

**Supplementary Table 1:** Characteristics of patients included in TA scRNAseq analyses of ARDS phenotypes from the COMET cohort. Normally distributed values are reported as mean ± SD. Non-normally distributed values are reported as median [IQR]. Categorical data are reported as N (% of total for category). P-values are for a t-test for normally distributed continuous data, Wilcoxon rank-sum for non-normally distributed, and chi-square test for categorical data.

|  | **Hyperinflammatory** | **Hypoinflammatory** |  | P |
| --- | --- | --- | --- | --- |
| **N** | 5 | 20 |  |  |
| **Age** | 62 [54, 73] | 63 [47, 71] |  | 1.00 |
| **Female** | 3 (60) | 9 (45) |  | 0.71 |
| **BMI (kg/m^2^)** | 25.5 [25.0, 30.0] | 28.1 [24.4, 38.0] |  | 0.68 |
| **Vasopressors at enrollment** | 4 (80) | 9 (45) |  | 0.37 |
| **Minimum PF ratio (mmHg)** | 74 [61, 92] | 108 [68, 135] |  | 0.16 |
| **SOFA score at enrollment** | 19 [15, 19] | 9 [7, 11] |  | <0.01 |
| **IL-8, pg/ml** | 236 [121, 424] | 12 [9, 25] |  | <0.01 |
| **Protein C, % control** | 54 [31, 56] | 104 [69, 132] |  | 0.01 |
| **Immunosuppression** | 1 (20) | 1 (5) |  | 0.85 |
| **Primary ALI Risk Factor** |  |  |  | 0.42 |
| *Pneumonia* | 2 (40) | 11 (56) |  |  |
| *Sepsis* | 2 (40) | 2 (13) |  |  |
| *Aspiration* | 1 (20) | 6 (25) |  |  |
| *None* | 0 (0) | 1 (3) |  |  |
| **Clinical microbiology** |  |  |  |  |
| *Respiratory viral pathogen* | 1 (20) | 1 (0) |  | 0.85 |
| *Respiratory bacterial pathogen* | 2 (40) | 7 (31) |  | 1.00 |
| *Extrapulmonary virus* | 0 (0) | 1 (5) |  | 1.00 |
| *Extrapulmonary bacteria* | 1 (20) | 1 (6) |  | 0.85 |
| *Extrapulmonary fungal infection* | 2 (40) | 0 (0) |  | 0.04 |

**Supplementary Table 2:** Characteristics of patients included in OLink Proteomic analysis of ARDS phenotypes from the Acute Lung Injury in Critical Illness cohort. Normally distributed values are reported as mean ± SD. Non-normally distributed values are reported as median [IQR]. Categorical data are reported as N (% of total for category). P-values are for a t-test for normally distributed continuous data, Wilcoxon rank-sum for non-normally distributed, and chi-square test for categorical data
