## Supplementary figures and images for "Distinct respiratory tract biological pathways characterizing ARDS molecular phenotypes"

### Supplementary Figure S1

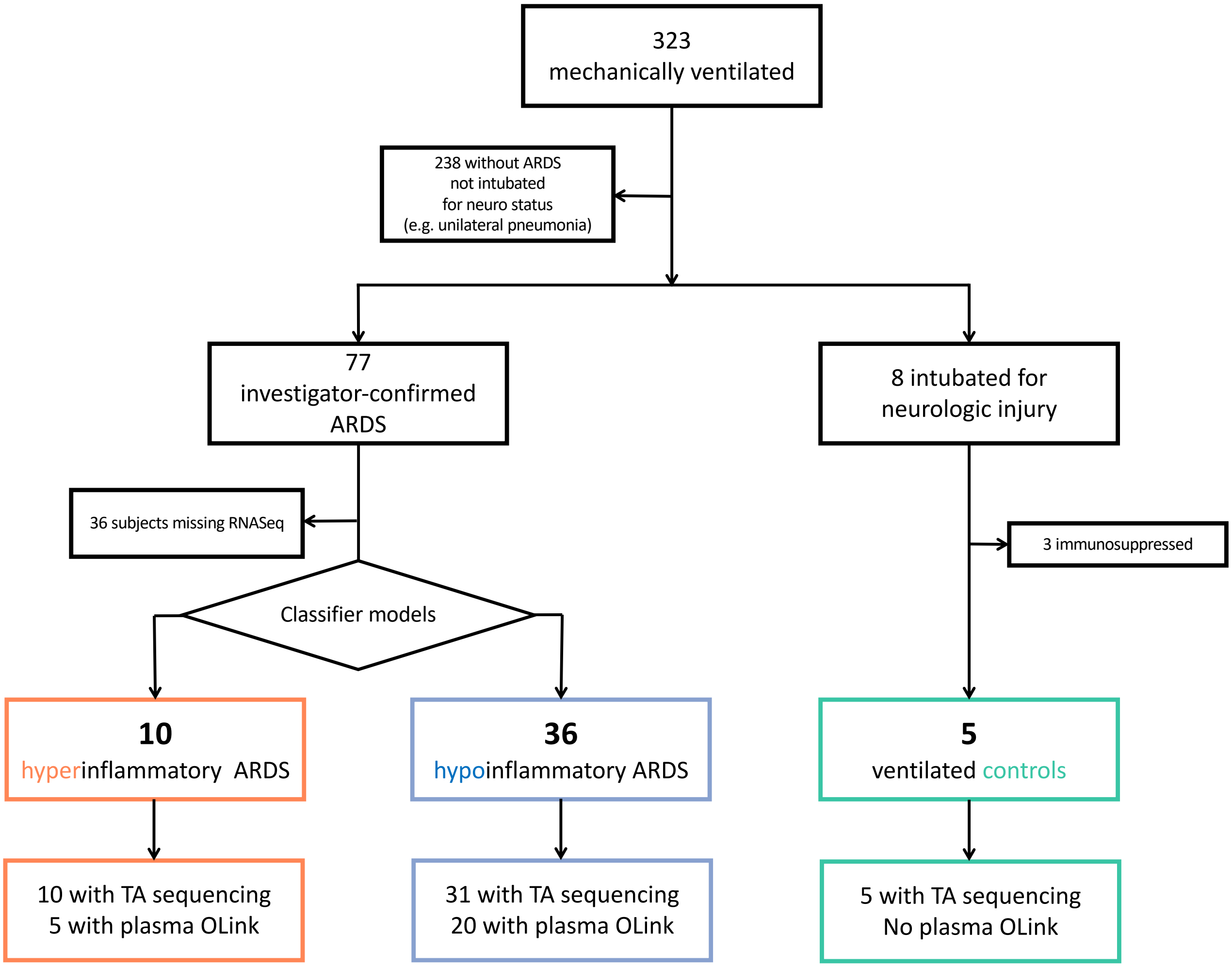

### Supplementary Figure S2

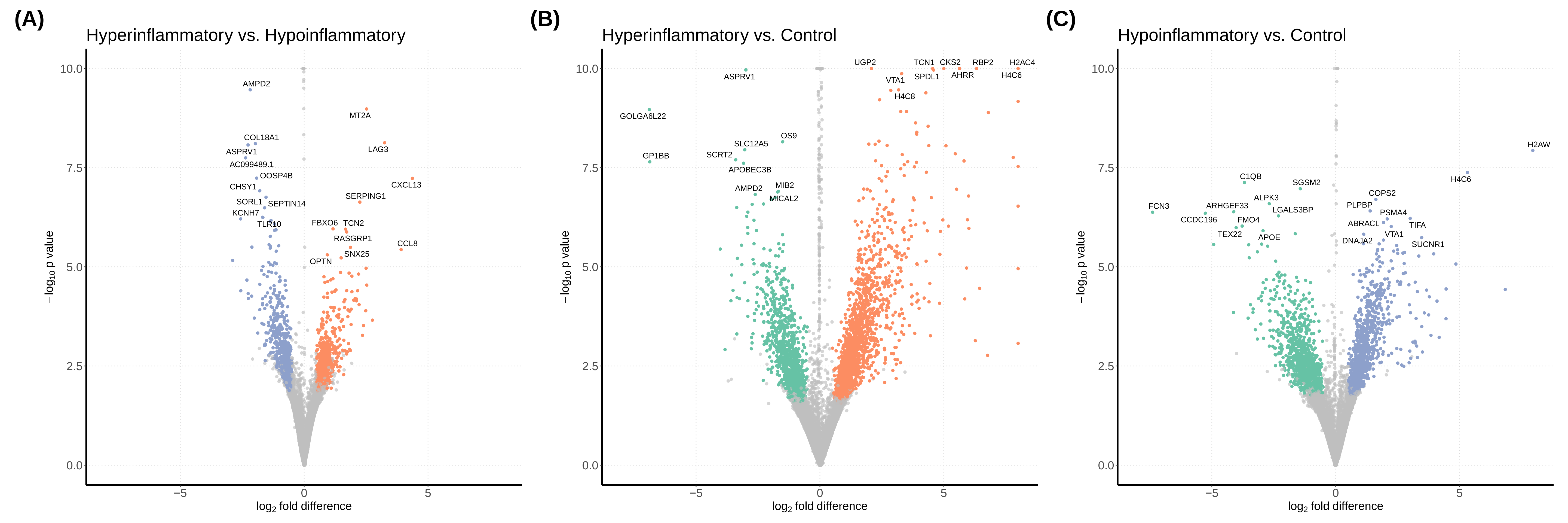

### Supplementary Figure S3

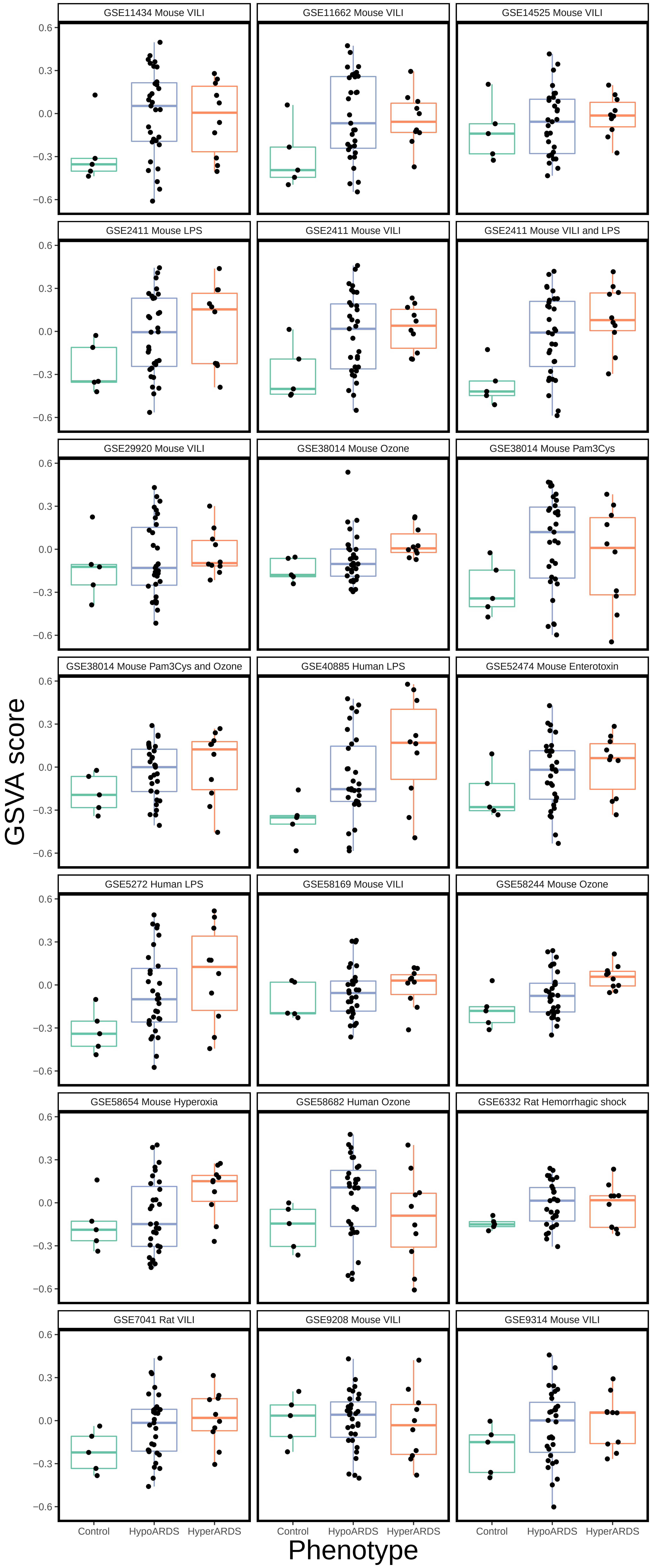

### Supplementary Figure S4

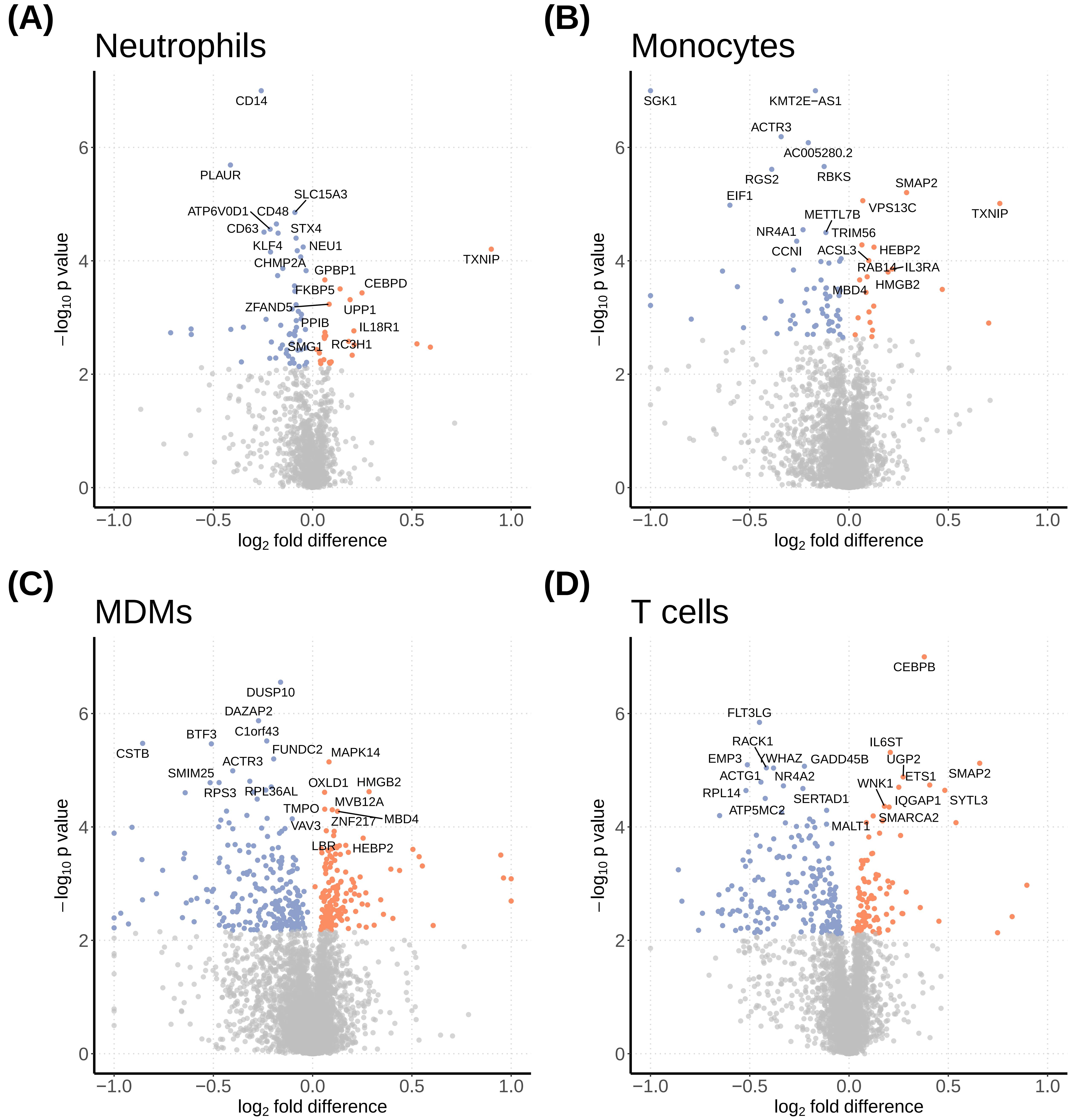

### Supplementary Figure S5

**(A)**

Upregulated signaling in HyperARDS

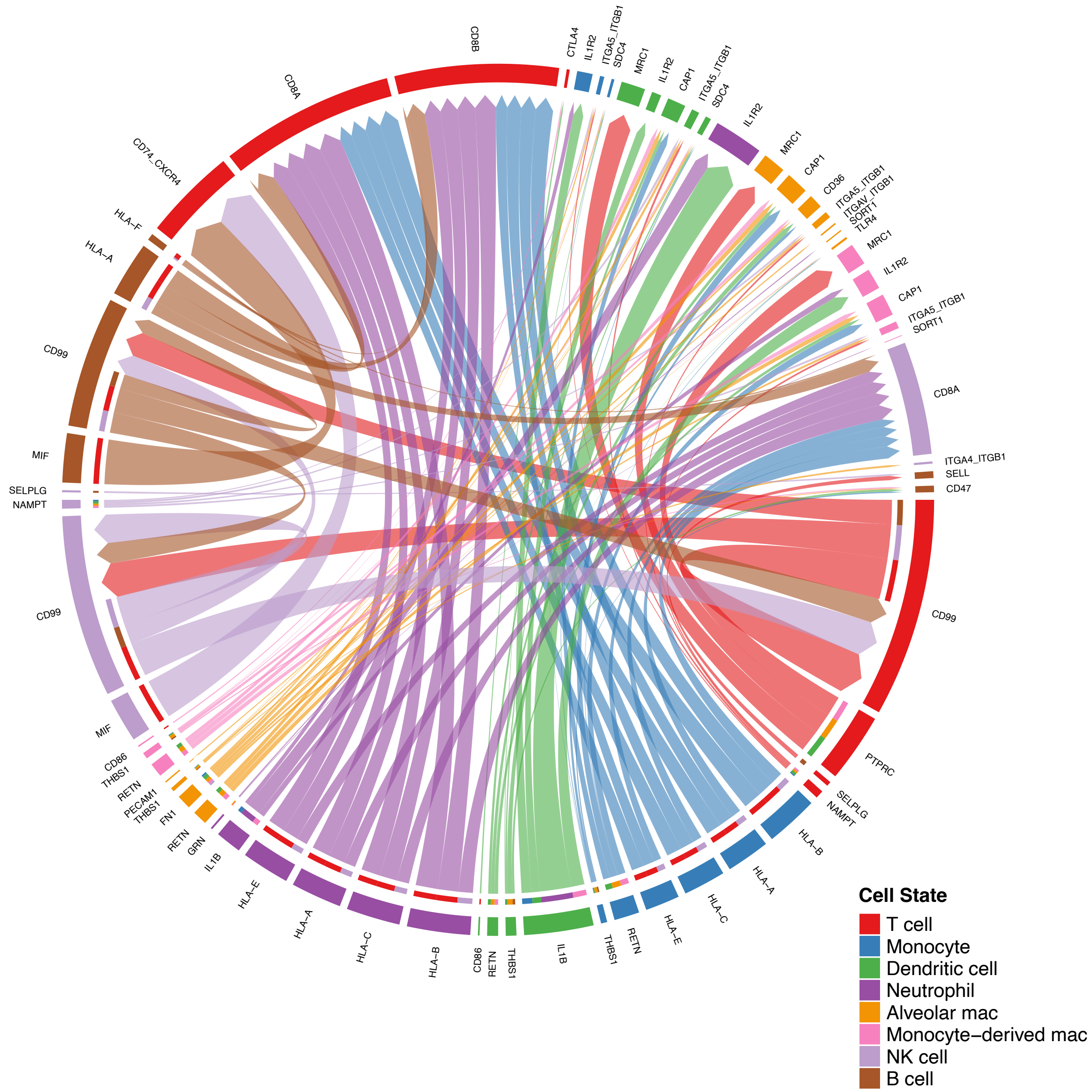

**(B)**

Upregulated signaling in HypoARDS

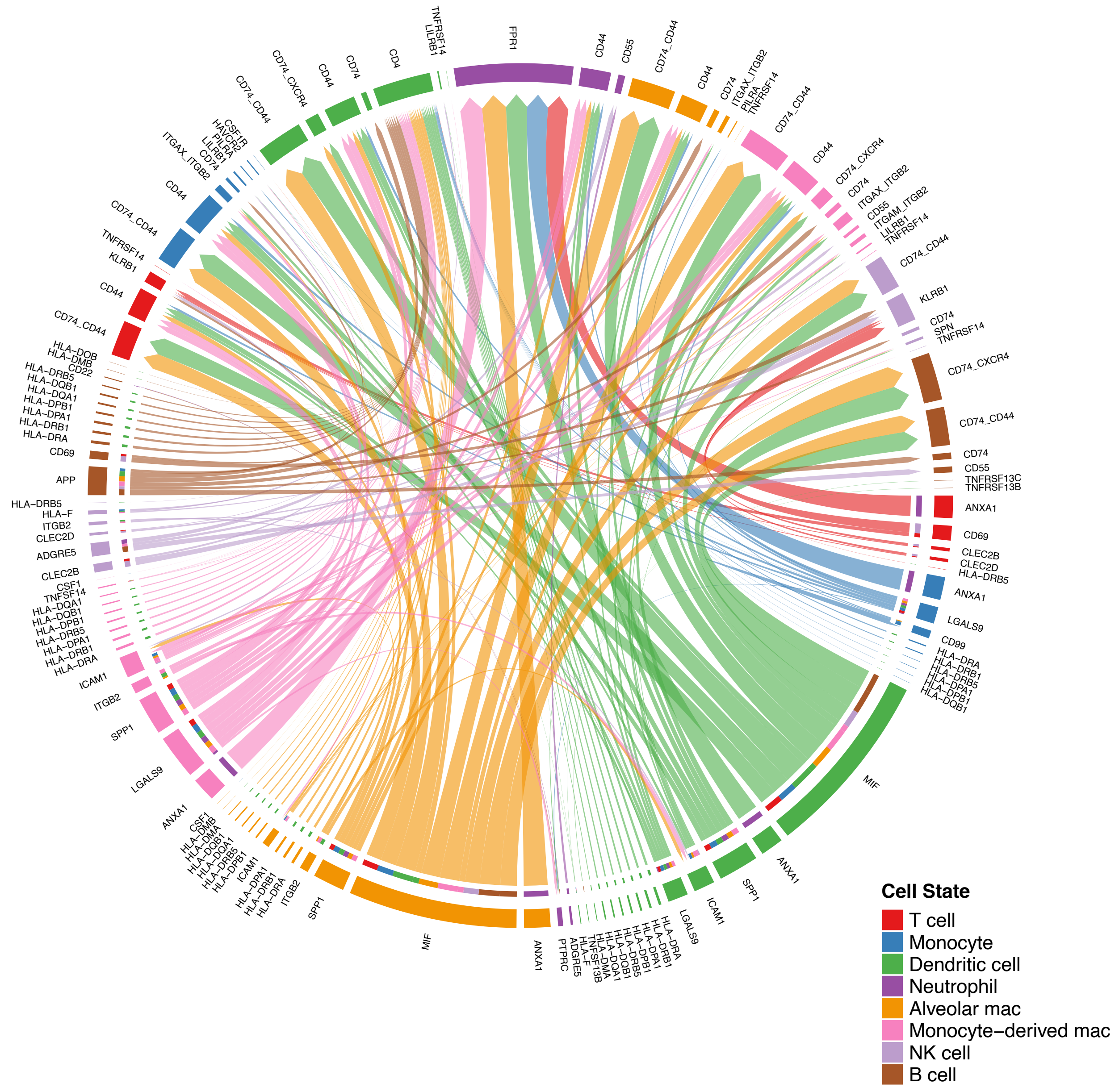

### Supplementary Figure S6

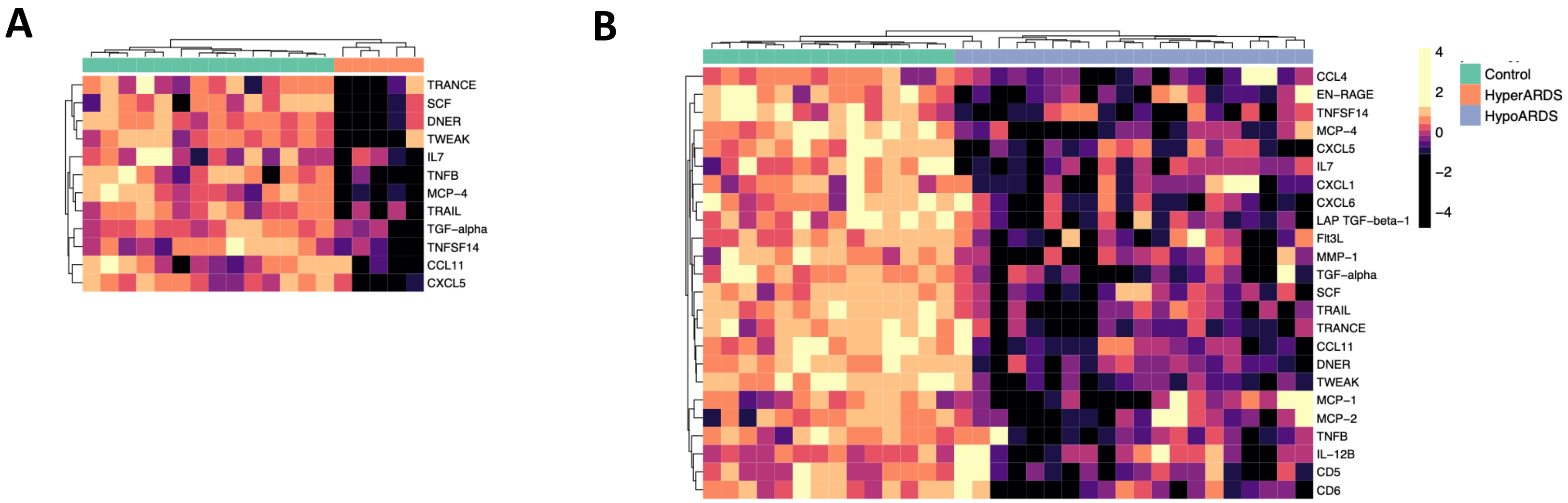

### Supplementary Figure S7

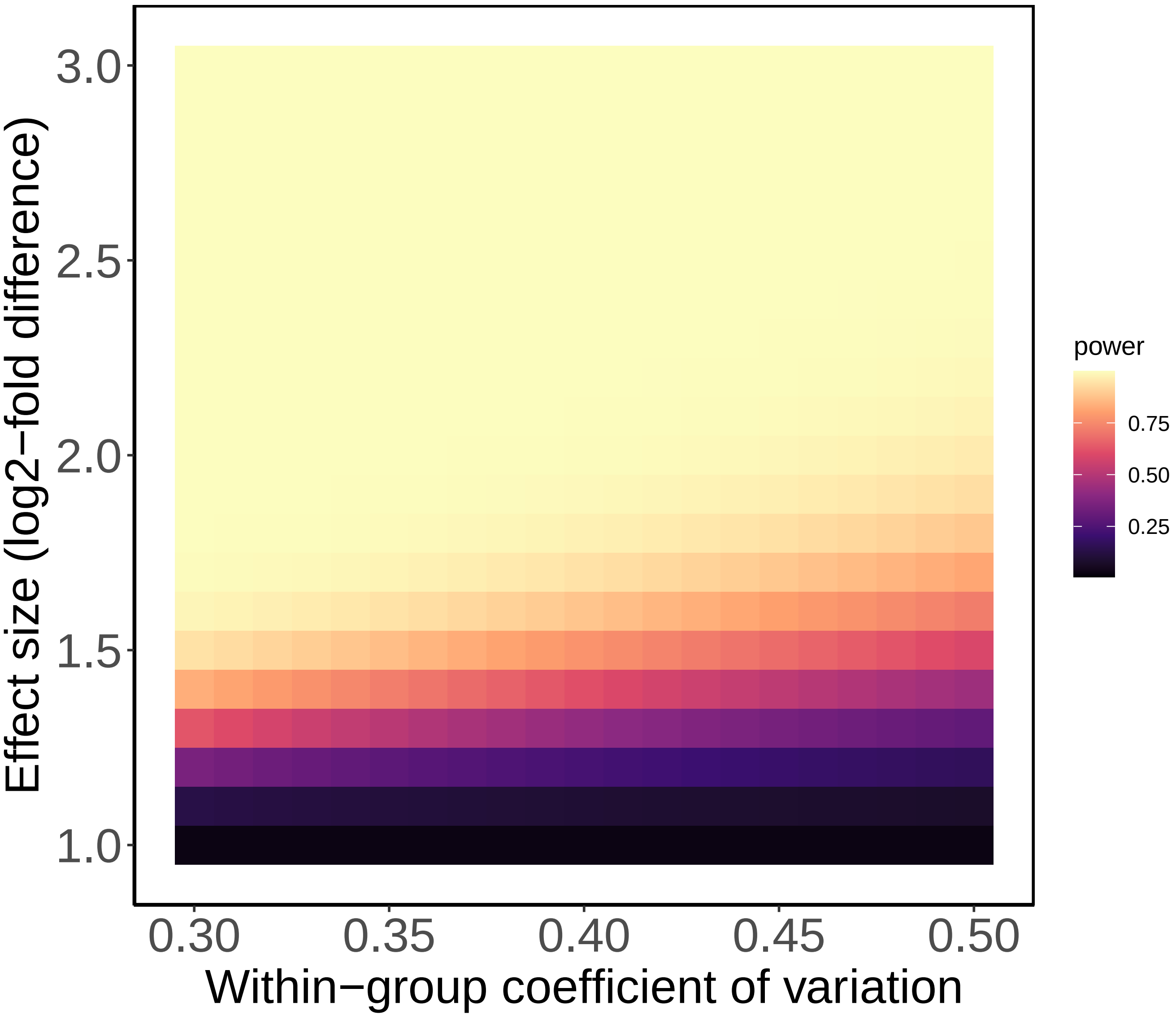
